# Development of a microcosting protocol to determine the economic cost of diagnostic genomic testing for rare diseases in Australia

**DOI:** 10.1101/2023.09.04.23295012

**Authors:** Dylan A Mordaunt, Zornitza Stark, Francisco Santos Gonzalez, Kim Dalziel, Ilias Goranitis

## Abstract

**Introduction:** Genomic testing is a relatively new, disruptive and complex health technology with multiple clinical applications in rare diseases, cancer and infection control. Genomic testing is increasingly being implemented into clinical practice, following regulatory approval, funding and adoption in models of care, particularly in the area of rare disease diagnosis. A significant barrier to the adoption and implementation of genomic testing is funding. What remains unclear is what the cost of genomic testing is, what the underlying drivers of cost are and whether policy differences contribute to cost variance in different jurisdictions, such as the requirement to have staff with a medical license involved in testing. This costing study will be useful in future economic evaluations and health technology assessments to inform optimal levels of reimbursement and to support comprehensive and comparable assessment of healthcare resource utilisation in the delivery of genomic testing globally.

**Methods:** A framework is presented that focuses on uncovering the process of genomic testing for any given laboratory, evaluating its utilization and unit costs, and modelling the cost drivers and overall expenses associated with delivering genomic testing. The goal is to aid in refining and implementing policies regarding both the regulation and funding of genomic testing. A process-focused (activity-based) methodology is outlined, which encompasses resources, assesses individual cost components through a combination of bottom-up and top-down microcosting techniques and allows disaggregation of resource type and process step.

**Ethics and dissemination:** The outputs of the study will be reported a relevant regional genetics and health economics conferences, as well as submitted to a peer-reviewed journal focusing on genomics.

**Article Summary:**

- This method uses key stakeholder interviews, health care resource utilisation and unit cost data collection for estimating the economic cost of diagnostic genomic testing in Australia.
- Advancements may include time in motion studies or greater reporting detail but with a trade-off in feasibility.
- Probabilistic modelling addresses uncertainty inherent in input estimates.
- The study does not incorporate variations in testing pathways, such as resequencing stored DNA costs or reanalysis of existing sequencing data.
- Automating ongoing monitoring of changes in genomic testing workflows, resource utilisation and costs would be beneficial.

## Introduction

Genomic testing holds the potential to transform diagnosis and health outcomes in rare diseases at scale (1-3). Genomic testing is increasingly offered to individuals with rare diseases, particularly in childhood, as a first-line test (4). Over the past decade, reimbursement for genetic testing with chromosomal microarray has added a small increment to the yield of the diagnostic workup in individuals with rare disorders (5). Translational and clinical research studies have demonstrated substantial increases in diagnostic yield associated with diagnostic genomics approaches that utilise massively parallel sequencing (MPS) (6-8). Genomic data generation, analytics, interpretation and clinical workflows vary widely. These variations have implications for resource utilisation and the corresponding cost of rare disease diagnosis.

Public reimbursement for rare disease genomic testing began in Australia in May 2020 for syndromic and non-syndromic intellectual disability (9). However, the optimum reimbursement level is yet to be determined, as are the costs related to testing for a wider range of conditions. Micro-costing studies have shown promise for more precise cost estimations for a small subset of genomic test indications (10-13). However, differing patient cohorts and workflows, and intra-jurisdictional variation are likely to contribute to significant heterogeneity in genomic test costs (14), affecting the generalisability of micro-costing estimations.

Microcosting studies published to date come from small-scale single centres, and there is limited information from clinical centres working at scale, and in a comparative manner. Several studies have explored the cost*-effectiveness* associated with rare disease genomic testing, but do not include a detailed or microcosting component (15).

Recently, a study by Gonzalez, et al. (16) systematically reviewed microcosting studies of genomic testing finding a range of costs estimates between US$2,094-$9,706 and US$716-$4,817 per patient, for genome sequencing and exome sequencing, respectively. Seven studies were identified from 2016 to 2022, with study contexts varying from research through clinical. There are varying challenges with the studies identified in this systematic review, ranging from reproducibility, the completeness of the information, gaps in certain costs (for example bioinformatics, physical and data storage), and being limited to genomics applied to specific scenarios (e.g. intellectual disability and cardiac conditions). Many of the studies are now several years old and given that there appears to be temporal changes in costs, warrant updating.

Microcosting is a method used to disaggregate complex technologies to better understand the source and magnitude of resource use components and total costs. In the example of a complex biomedical technology like genomic sequencing, the outputs of sequencing combine expensive and specialised equipment, consumables and highly skilled professionals in the context of widely varying regulatory environments and reimbursement. The disaggregated nature of the technology combined with the rapidly changing pace of component technologies, add additional complexity to the operation of genomic technologies (17).

Microcosting studies have several strengths, including that they provide detailed cost information, comparability, and flexibility (14 18 19). By breaking down costs into individual components, microcosting studies provide in-depth insights into the resources used and the expenses incurred in the delivery of diagnostic genomic testing. Microcosting studies enable consistency in the comparisons between different testing approaches, healthcare settings, and countries, informing decision-making and policy development. They can be adapted to a wide range of genomic testing scenarios and rare diseases, allowing for an accurate and comparable assessment of resource utilisation and associated costs in diverse contexts.

## Methods

These methods will be developed and presented according to the reference case guideline and checklist developed by the Global Health Cost Consortium (20).

### Aims and Objectives

The objective of this study is to undertake microcosting of rare disease genomic testing in the Australian environment to add to the global understanding of the cost of genomic testing. This study will explore various policy questions, including the optimum reimbursement rates for rare disease genomic testing. Specifically, we aim to cost genomic testing based on testing undertaken by Australian Genomics first phase flagship studies and refers to testing using genome-wide sequencing approaches such as whole exome sequencing, whole genome sequencing, gene panels or variants such as clinical exomes. Testing context is inclusive of singleton and trio testing.

The data collected will be used to inform public policy as it relates to reimbursement both in Australasia and in a global context. The intention is to present a method that presents disaggregated steps in a process and resource utilisation so that these can be modelled to understand component costs, and sources of variability and for these to be comparable to other jurisdictions nationally and internationally. The information will also be used to explore policy settings and to understand drivers of genomic testing costs globally as well as in Australia. This microcosting analysis will be used to help understand whether current reimbursement levels for rare disease genomics are adequate in Australia, particularly through the Medicare Benefits Services (MBS) scheme, and to inform health economic evaluations. The primary aim of this study is to determine the economic cost of rare disease genomic testing in Australia and the key cost drivers. Secondary aims include determining whether there is jurisdictional cost variance, and what the drivers of cost variance are. Information will be presented in a way that is generalisable across Australia and reproducible and transferable to global contexts.

### Study Design

A microcosting study will be undertaken using mixed top-down and bottom-up approaches to identifying healthcare resource utilisation and associated unit costs. Top-down microcosting entails the allocation of resources and costs based on existing aggregate data, as opposed to collecting detailed, individual-level data (which would be characteristic of a bottom-up microcosting approach). Top-down microcosting will be used where newly disaggregated data is not able to be collected.

Process mapping will be utilised to identify the steps in the process. Expert interviews will be used to collect information on resource utilisation, as will laboratory and computer logs, as well as accounting data sources. List prices will be obtained from the laboratory, where available, and substituted by vendor pricing where not. Deterministic sensitivity analysis will be undertaken on the utilisation of large instruments such as sequencers and robotics to understand the effect of changing single microcosting-related parameters on the overall cost of genomic testing. Probabilistic sensitivity analysis will be undertaken on resource utilisation and unit costs to incorporate inherent uncertainty in costing inputs on the cost estimate.

### Intervention

Rare disease genomic testing is a technology involving massively parallel sequencing (MPS) of germline DNA to assist in the timely diagnosis of rare diseases. The target population of this technology are children and adults with undiagnosed diseases, and specifically those without a known molecular cause of their diagnosis. In practice, this is a broad group. For this study, we focused on the rare disease testing indications evaluated as part of Australian Genomics flagship projects (21),(22) between 2016 and 2021. Australian Genomics is a national genomic medicine initiative, which was funded to accelerate the implementation of genomics into healthcare. The cohorts were recruited prospectively and underwent genomic testing largely through clinically accredited laboratories in Australia. Evaluation of the diagnostic and clinical utility of genomic testing in these cohorts, as well as cost-effectiveness, has served as the basis for the application to publicly fund genomic testing across a range of clinical indications. In addition to MBS funding for syndromic and non-syndromic intellectual disability in 2020, genomic testing for a range of cardiac and renal genetic disorders has more recently become available in mid-2022, with many more applications in progress.

Seven Australian diagnostic laboratories are accredited to provide rare disease genomic testing. These laboratory services serve as state or supra-regional referral laboratories from local health services. Each laboratory is primarily based in a single facility, though samples are sent from a wide range of health services.

Each “genomic test” will be described in detail in the results section since disaggregation of the sequencing procedure, and underlying process to produce the test result is a core part of this study’s process-centred microcosting method. In general, the process involved in a genomic test comprises the receipt of a tissue or fluid sample, processing to extract DNA and subsequent storage of DNA, the preparation of the DNA for sequencing, sequencing of DNA, assembly and analysis of data, quality assessment of sequencing and resulting data, interpretation of data, reporting of results, oversight and issuance of results. Several indirect processes contribute to results production, including the development and maintenance of the genomics assay and bioinformatics pipeline.

### Economic perspective taken

In this study protocol on microcosting, we adopt the economic perspective of the laboratory provider, taking into account local, jurisdictional, and federal health system components that influence laboratory sustainability, health provider access, and consumer access. Our goal is to determine the full real-world economic cost to the provider of operating a genomics laboratory, net of future costs, considering the multi-year lifespan of relevant component equipment.

As we aim to identify cost variation across jurisdictions, we will disaggregate different resourcing arrangements, including flat infrastructure and energy fees. The direct cost of producing a rare disease genomic test result will serve as the ultimate unit cost product in this exercise.

Although assessing the quality of service in genomics is complex, a standards-based approach is applied to diagnostic laboratory testing. Quality-of-service policies in Australia are extensive and include the National Association of Testing Authorities (NATA), International Association of Testing Authorities (IATA), International Organization for Standardization (ISO) (23 24), National Pathology Accreditation Advisory Council (NPAAC), and The Australian Council on Healthcare Standards (ACHS) standards related to diagnostic laboratories and clinical genomic testing (25). Genomic testing in Australia generally excludes companion pharmacogenomic testing and reporting of secondary findings.

Given the rapid evolution of genomics technology, we consider the acquisition, implementation, maintenance, and de-implementation of component costs involved in producing the genomic test as part of our costing exercise. Component costs are disaggregated according to the investment timeframe for determining net present value and from the time of acquisition, using 2022 as the base price year. The cost estimation excludes general laboratory start-up expenses, such as accreditation costs, but includes genomic testing start-up costs, such as instrument acquisition.

### Resource Use Measurement

The scope of inputs includes direct genomic testing service delivery costs, including physical biospecimen and data storage costs. Information on resource utilisation will be incorporated into the unit cost calculation. Indirect costs will be collected where possible and assumptions will be documented about overhead costs where they are not-for example, power usage is unlikely to be directly measured, but incorporated into overheads since it is anticipated to be a considerable component of instrument and computational operation costs. Costs required to start up, implement or maintain the unit’s production will be considered direct costs, including supporting changes within the production process. Test development costs, such as the development of the bioinformatics pipeline, will be included if they are directly related to the work-up of changes in the production of the unit (i.e. directly related to the development of the clinical genomic test), however, given the close relationship of this environment to research, research costs involved in test development will not be exhaustively collected. Corporate services costs, such as payroll and financial management, will be considered indirect, and not collected. Genomic testing costs included are outlined in Table 1. Costs categories.

**Table 1.** Costs categories.

| Category | Outcomes Estimated |
| --- | --- |
| Labour | Personnel classification - e.g. technician |
|  | Utilisation - minutes<br>Unit cost - i.e. salary / wage information |
| Equipment | Equipment type and vendor<br>Utilisation – instrument - total sample throughput<br>Utilisation – compute – nodes, CPUs/cores used, RAM used, duration of usage<br>Unit cost (e.g. cost of instrument, maintenance contracts, shipping, installation and configuration) |
| Consumables | Consumable type and vendor<br>Utilisation (volume, count or fraction of total unit)<br>Unit cost (exclusive of discounts) |
| Storage | Storage type (e.g. fridge) and vendor<br>Utilisation – total samples per storage type<br>Unit cost of storage unit<br>Computer storage type (e.g. fast storage, archival storage)<br>Utilisation – space utilisation on storage type<br>Unit cost of computer storage unit |

Estimation of the inputs will be by measurement of inputs using bottom-up microcosting for labour and consumables and top-down microcosting for equipment (26). The method of allocation of human resources, equipment and consumable inputs will follow semi-structured interviews with laboratory management (26). The semi-structured approach will involve determining the major components of the process undertaken to produce the costs (i.e. process mapping) (27), then identifying the time or amount utilised in each component, followed by determining the cost of each of the resources utilised. For labour costs these will be costed using standard pay scales (in the Australian environment determined by enterprise bargaining agreements, EBAs), identifying a suitable setting for the grade used within each classification. For equipment and consumable costs, the inputs will be collected by contacting the relevant vendor and obtaining list/market prices. Where required, the manufacturer or price will be kept confidential and redacted in the manuscript.

Representative utilisation of the production environment for producing each type of unit will be obtained from the laboratory during the interviews. This utilisation represented actual recorded utilisation rather than expert opinion on the approximate utilisation. Total utilisation therefore reflected the actual total utilisation for rare disease genomics in this laboratory, for the 12 months before the interviews.

Interviews will be conducted with a small number of senior scientists directly involved in designing, operating, managing and troubleshooting the production pipeline. Resource utilisation will be obtained from internal spreadsheets accounting for resource utilisation (equipment and consumables), and internally performed time-in-motion studies for labour utilisation. Where there will be missing data, either assumptions will be made or inputs will be sought from the literature, adjusted to Australian dollar value in 2022 and annotated in the model.

Data will be obtained in a period from May through August 2022, will be collected prospectively by the laboratory, longitudinal in nature but treated as cross-sectional for the purpose of this study. The recall period will be the 12 months prior to the study.

### Valuation

Unit cost data for labour use will be obtained through publicly available award and enterprise bargain agreements (26). Unit cost data for equipment and consumables will be obtained by contacting the relevant vendor and negotiating either identified or confidential access to market/list prices. Presentation of this information is redacted or treated as confidential accordingly in this manuscript. It is worth noting that local (Australian prices) for many of these are calculated based in US reference prices, adjusted for currency with substantial transport costs. Within the study period there has been a significant increase in cost of some of these.

The approach to amortisation will be straight-line, utilising Australian Taxation Office (ATO) rates for 2022 rather than state or local government policies (28 29). Residual value will be considered to be zero at the end of the assessment period. The rationale for this is to allow comparison between jurisdictions. Life years for each of the instruments/equipment is reported individually. Discount, inflation and conversion rates – Benchmark interest rates from the Australian Taxation Office will be used for discounting, inflation and conversion (28), that rate for 2022 is 4.52%. Net present value will be also adjusted for inflation using the ATO rate for CPI (29).

None of the component costs required shadow pricing. Although in some centre’s genomic testing pipelines, there are regular case review meetings and wide consultation, the assumption will be made that all of the needed labour will be incorporated into the costing model. Whilst occasionally experts outside of the service are consulted informally for advice, these shadow resources will not be incorporated.

The aforementioned method will be repeated for each of the study sites, in an attempt to analyse variation. Component fixed and variable costs are aggregated and analysed both in terms of total resource sub-group costs and total costs for component of the pipeline.

### Patient and Public Involvement

This study explores a laboratory testing method from which the consumer/patient usually only sees one of four elements-1) collection of the specimen such as by venepuncture, 2) the order form, 3) an invoice for the cost to the patient of the test (in contexts where consumers pay directly for this test) and 4) the outputs (report) from the test. In many respects the method seeks to systematically understand the outputs. However, the study does not include patient, consumer, or public involvement. The participants (stakeholders) in the study are workers in the clinical laboratory. Results will be disseminated to these workers.

### Understanding Checking

Following the iterative interviewing process and collecting of data, a process will be developed and associated input tables to reflect the resources used throughout the process. These tables will be represented to key stakeholders from the laboratory to clarify understanding of the process, the resources utilised and to help to refine any estimates of inputs.

### Modelling of Outputs

Outputs will be modelled leveraging the inputs entered into the model using Microsoft Excel and Stata. These outputs will be reported by category of resource and step of the process. Deterministic sensitivity analysis will be undertaken to explore the cost impact of plausible variations in key assumptions and parameter values-the main equipment utilisation for consideration is likely to be instrument usage such as sequencer and robotics (13). Probabilistic sensitivity analysis will be undertaken on model inputs, to incorporate uncertainty in items where either large input variance is anticipated based on the interviews, or where the nature of the input is clearly a broad estimate (e.g. expert estimates) (26). The characteristics of the input distribution will be based on information collected in the interviews. These will then be used to a generate a full probabilistic analysis of the model, to strengthen the transferability and generalisability of the model based on the inputs and make probabilistic estimates of the certainty of the outputs (30-32).

### Strengths and Limitations

Despite their strengths, microcosting studies also have limitations, particularly generalisability, data availability and quality, and complexity (14 18 19). The findings from microcosting studies may not be directly applicable to other settings, given the variability in costs and resource use across different healthcare systems and countries. The accuracy and completeness of cost data can vary, potentially affecting the reliability of cost estimates. Microcosting studies can be resource-intensive and time-consuming, given the level of detail required to capture all cost components.

This method leverages key stakeholder interviews within the laboratory who will make estimates of model inputs, as well as draw from other data sources such as utilisation and costing spreadsheets. These inputs may be directly measured in some cases, such as the measured volume of tests performed. However, for other inputs such as labour utilisation, these will be expert estimates. Advancement of the method could involve, for instance, time in motion studies or incrementally greater levels of detail. However, there is a trade-off in terms of incremental yield and feasibility of the study. Time-in-motion studies also produce errors, such as a Hawthorn effect. Hence probabilistic modelling will be used to measure the uncertainty inherent to the inputs. Resequencing of stored samples currently occurs, however the cost of resequencing stored DNA compared with newly collected specimens isn’t incorporated as most tests are currently on new samples. Reanalysis of existing sequencing data is a workflow that is accounted for in this study. Finally, continuous monitoring of genomic testing costs is important (as is monitoring of utilisation)-part of the narrative in genomic testing is not only of costs but changing costs in different contexts and with time. However, this study does not attempt to create a mechanism for ongoing monitoring of genomic testing costs.

We do not address the issue of general laboratory start-up costs in this study. Whilst scale and context of the laboratory are likely to influence the operation of a genomics laboratory, assessing the general laboratory start-up costs introduces additional variability and complexity, that is important but different in scope and focus. Specifically, infrastructure choices are likely to be interdependent on the other services offered by a laboratory. We believe the outputs of the study are likely to lend themselves to specific questions around scale, complexity, interdependent services and infrastructure, feeding into future research.

### Ethics and dissemination

The outputs of the study will be reported at relevant regional genetics and health economics conferences, as well as submitted to a peer reviewed journal focusing on genomics.

## Data Availability

n/a

## Acknowledgements

There are no additional acknowledgements for the development of the protocol.

## Data Statement

This protocol development did not collect data.

## Author Contributions

DM planned and executed the protocol development and wrote the manuscript. ZS, IG, KD conceived, planned and oversaw the protocol development, and oversaw the writing. FS contributed to the planning of the protocol development and writing of the manuscript.

